# Availability and readiness of healthcare facilities and their effects on under-five mortality in Bangladesh: Analysis of linked data

**DOI:** 10.1101/2022.06.22.22276753

**Authors:** Md. Nuruzzaman Khan, Nahidha Islam Trisha, Md. Mamunur Rashid

## Abstract

**Background:** Under-five mortality is unacceptably high in Bangladesh instead of governmental level efforts to reduce its prevalence over the years. Increased availability and access to the healthcare services can play a significant role to reduce under-five mortality. We explored the associations of several forms of child mortality with health facility level factors adjusted for individual-, household-, and community level factors.

**Methods:** The 2017-18 Bangladesh Demographic and Health Survey data and 2017 Bangladesh Health Facility Survey data were linked and analysed. Our outcome variables were neonatal mortality, infant mortality, and under-five mortality. Health facility level factors were considered as major explanatory variables. They were the basic management and administrative system of the healthcare facility, availability of the child healthcare services at the nearest healthcare facility, readiness of the nearest healthcare facility to provide child healthcare services and the average distance of the nearest healthcare facility providing child healthcare services. The associations between the outcome variables and explanatory variables were determined using the multilevel mixed-effect logistic regression model.

**Results:** Reported under-five, infant and neonatal mortality were 40, 27, and 22 per 10000 live births, respectively. The likelihood of neonatal mortality was found to be declined by 15% for every unit increase in the score of the basic management and administrative system of the mothers’ homes nearest healthcare facility where child healthcare services are available. Similarly, the availability and readiness of the mothers’ homes nearest healthcare facilities to provide child healthcare services were found to be linked with the 18-24% reduction in neonatal and infant mortality. On contrary, for every kilometre increased distance between mothers’ homes and their nearest healthcare facility was found to be associated with a 15-20% increase in the likelihoods of neonatal, infant and under-five mortality.

**Conclusion:** The availability of health facilities providing child healthcare services close to residence and their improved management, infrastructure, and readiness to provide child healthcare services play a significant role in reducing under-five mortality in Bangladesh. Policies and programs should prioritize to increase the availability and accessibility of health facilities that provide child healthcare services.

## Introduction

The Sustainable Development Goals (SDG) started in 2015 with ambitious targets to reduce the neonatal and under-five mortality rates to 12 and 25 per 1000 live births, respectively, by 2030 [1]. The Millennium Development Goals (MDGs), which ended in 2015 with more than 50% (from 90 to 43 per 1000 live births) reduction of global under-five mortality, helped the world to make such highly ambitious targets [2,3]. However, such an average estimate of under-five mortality as well as reduction during the MDGs does not truly represent the situation of under-five mortality in Low- and Middle-Income Countries (LMICs). The rate of under-five mortality in LMICs is still 69 per 1000 live birth-a rate that was the global average on the 2000s [4,5]. Consequently, LMICs represent more than 90% of the 5 million under-five deaths that occur globally, and the probability of under-five mortality is 14 times higher in LMICs than in developed countries [5].

The rate of under-five mortality is even higher in Asian and African regions where around 80% of the global under-five deaths occur, though these two regions account for only 53% of the global life births [6]. The causes of such higher under-five mortality in LMICs, in particular Asian and African regions, are infectious diseases including pneumonia, diarrhoea and malaria as well as pre-term birth complications, birth asphyxia and trauma, and congenital anomalies [5,7]. Cost-effective, proven interventions, including quality of maternal healthcare services around the time of birth and child healthcare services, exist to prevent and treat infectious diseases and other causes associated with under-five mortality [5]. This requires trained and equipped health workers, including midwives and the availability of essential commodities [6].

Bangladesh contributes around 4.30% of the total live births in the South Asian region with around 3 million yearly live births [8]. However, the country is ranked number 4 in terms of neonatal mortality in South Asia, whereas Pakistan, Afghanistan and India are ranked in the first three, respectively [5]. Such a higher rate of child mortality in Bangladesh is reported instead of its significant progress in preventing infectious diseases, including diarrhoea and pneumonia, increasing breastfeeding practice and immunization, improvement of maternal undernutrition, reduction in adolescent pregnancy, and a significant increase in maternal healthcare services use [8-11]. Similar progress has been reported in other South Asian countries as well as LMICs [12-15]. Consequently, most of the LMICs, including Bangladesh are currently off-track to achieving the SDGs targets related to child mortality [16,17].

Healthcare facilities in Bangladesh and LMICs are often criticized for the poor quality of available services, and lack of healthcare personnel and equipment [11]. However, a significant progress has been achieved over the decades, particularly during the MDGs [2]. The pace of progress is even now much faster than before to ensure the universal health coverage in line with the SDGs’ target [18]. Therefore, it is desirable that such progress is now making a significant contribution to reducing child mortality, though research to justify this understanding is lacking in Bangladesh and other LMICs. The reason for such lacking is the absence of longitudinal data as well as linked individual and healthcare facility level data. Consequently, available research mainly explored the socio-demographic and behavioural factors associated with child mortality, including maternal age and education, under-nutrition, and poor feeding practice where effects of healthcare facility level factors were ignored [19-23]. As the occurrence of child mortality is interconnected with multidimensional factors, including factors at individual-, household-, community-, and healthcare-facility level factors, therefore, determining such associations without considering the healthcare facility level factors may produce imprecise findings.

To address these limitations, we conducted this study to explore the association between healthcare facility level factors and child mortality in Bangladesh adjusting for individual-, household-, and community level factors.

## Methods

### Study design and sample

The 2017/18 Bangladesh Demographic and Health Survey data and 2017 Healthcare Facility Survey were linked and analysed in this study. Details of sampling procedures of these surveys are available in the respective survey reports [24,25].

The 2017/18 BDHS is the nationally representative household survey conducted as part of the Demographic and Health Survey Program of the USA. The survey used a validated questionnaire to collect data and received ethical approvals from the ICF International, USA and the Ministry of Health and Family Welfare of Bangladesh. The National Institute of Population Research and Training (NIPORT) conducted this survey along with the Mitra and Associates (a private research firm). The survey selected nationally representative households through two-stage stratified random sampling methods. At the first stage of the sampling, the survey selected a list of 675 clusters, covering each division and urban/rural area of Bangladesh. The clusters were selected from a list of 293,579 clusters of Bangladesh which was generated by the Bangladesh Bureau of Statistics as part of the 2011 Bangladesh National Population Census. A total of 672 clusters was retained finally after excluding 3 clusters because of the extreme flood. A fixed number of 30 households was selected at the second stage from each selected cluster. Finally, the survey was conducted in 19,457 households with an over 96% inclusion rate. There were 20,376 women in these selected households, of them, 20,127 women were interviewed with a response rate of 98.8%. Informed written consent was obtained from all participants [25].

The 2017 BHFS was also conducted as part of the Demography and Health Survey program of the USA. The survey collected data from 1,524 healthcare facilities selected proportionately from the public, private and non-government sectors. Both census and stratified random sampling methods were used to select these facilities from the 19,811 registered healthcare facilities in Bangladesh.

The GPS point locations are available for each of the 672 PSUs included in the 2017 BDHS and 1,524 healthcare facilities included in the 2017 BHFS. We linked the PSUs with the nearest healthcare facilities using the administrative boundary linkage method. The details of this method have been published elsewhere [26].

### Analytical sample

Conditions applied to be included in this study were: (i) the women who were given at least one live birth within five years preceding the survey and (ii) recorded survival status of the corresponding children. A total of 8,759 women met these criteria, as such they were analysed in this study.

### Outcome variables

We considered three outcome variables: neonatal mortality (death occurred within 1 month of live birth), infant mortality (deaths occurred within 12 months of live birth) and under-five mortality (death occurred within 60 months of live births). The BDHS recorded these mortality data by asking women whether they had any live birth within five years of the survey date and survival status of their respective children. In the occurrence of more than one live birth within five years, survival status data were collected for every child. These data were then recategorized by following the relevant guidelines of the WHO to generate the outcome variables [27].

### Explanatory variables

Health facility level factors were our primary explanatory variables. Several health facility level factors were considered, including availability of child healthcare services in the nearest healthcare facilities, readiness of the nearest healthcare facility to provide child healthcare services and average distance on road communication from mother’s resided cluster to the nearest healthcare facility where child healthcare services are available. The variables used to generate scores for these corresponding variables are presented in the Supplementary Table 1. A total of seven variables was used to generate score for the variable “ basic management and administrative system”. We considered 35 variables to generate the score for availability of child healthcare services in the nearest healthcare facilities. Total of 10 variables were considered to generate the corresponding score for the readiness of the nearest healthcare facility to provide child healthcare services. For generating scores, we have given “ 1” point if the service referred by that particular variable was available in the healthcare facility and “ 0” was given for non-availability. The combined score for each item considered under the particular dimension was then generated by adding the corresponding individual score. The average distance on-road communication of the nearest facility providing child healthcare services was also calculated and included as a health facility-level variable. The cluster nearest child healthcare facility was identified first. Then using Bangladesh road communication data, the average distance from the cluster to its nearest health facility was calculated [28].

Other explanatory factors were selected in two stages. At first, we generated a list of all possible confounders by reviewing the relevant studies conducted in LMICs [4,5,20,21,29-32]. Availability of the selected variables was then checked in the dataset we analysed. The variables available in the dataset were then considered together with considered healthcare facility level factors and forward regression models were run. The variables found insignificant at 0.20 or more were deleted. Multicollinearity was also checked. Factors that were finally selected were then categorized as individual-, household-, and community level factors and included in this study. Individual level factors were maternal age at birth, maternal education, and maternal formal employment status. Households level factors were partner’s educational status, sex of the household’s head, total children ever born, exposure to mass media, and wealth quintile. Place of residence and region of residence were considered as community level factors.

### Statistical analysis

Descriptive statistics were used to summarise the characteristics of the respondents. Multilevel mixed-effect logistic regression model was used to assess the associations between child mortality and health facility level factors adjusted for individual-, household-, and community-level factors. The reason for using the multilevel mixed-effect logistic regression model was hierarchical structure of the BDHS data, in which individuals are nested within a household and households are nested within a cluster. This creates multiple dependencies in the data for which multilevel mixed-effect logistic regression is deemed the appropriate approach [33]. Both unadjusted and adjusted models were run. Results are reported as Odds Ratios (OR) with 95% Confidence Intervals (95% CI). Statistical analyses were conducted using Statistical package R (version 4.10).

## Results

Background characteristics of the respondents are presented in Table 1. Mean age of the respondents was 24 years and mean years of education was 6.7 years. Mean age of the children was 29.1 months. Around 48% of the total under-five children were girls.

**Table 1:**
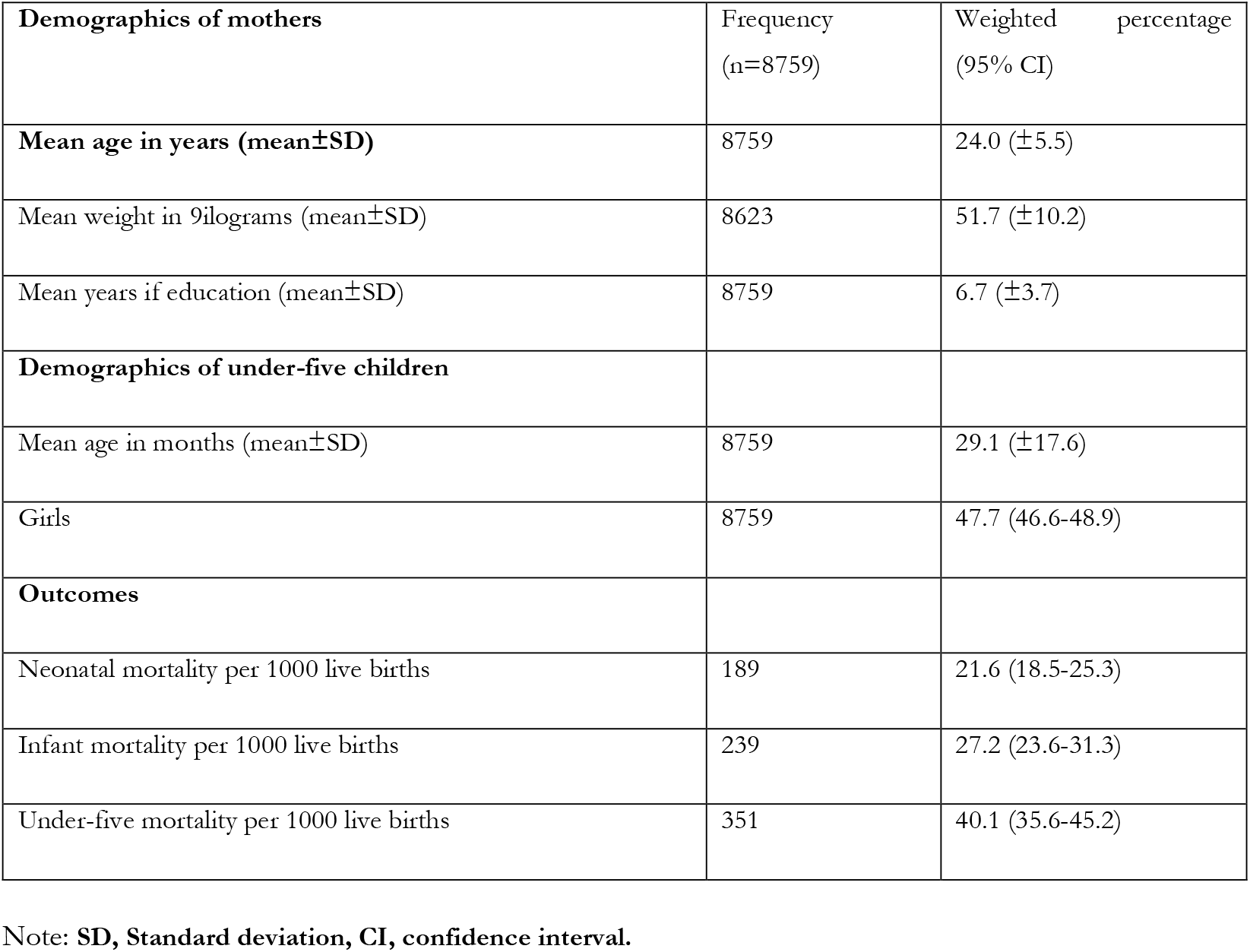
Key information about the study participants and outcome variables.

| **Demographics of mothers** | Frequency<br>(n=8759) | Weighted percentage<br>(95% CI) |
| --- | --- | --- |
| **Mean age in years (mean±SD)** | 8759 | 24.0 (±5.5) |
| Mean weight in kilograms (mean±SD) | 8623 | 51.7 (±10.2) |
| Mean years of education (mean±SD) | 8759 | 6.7 (±3.7) |
| **Demographics of under-five children** |  |  |
| Mean age in months (mean±SD) | 8759 | 29.1 (±17.6) |
| Girls | 8759 | 47.7 (46.6-48.9) |
| **Outcomes** |  |  |
| Neonatal mortality per 1000 live births | 189 | 21.6 (18.5-25.3) |
| Infant mortality per 1000 live births | 239 | 27.2 (23.6-31.3) |
| Under-five mortality per 1000 live births | 351 | 40.1 (35.6-45.2) |

Note: **SD**, Standard deviation, **CI**, confidence interval.

We explored the distribution of neonatal, infant and under-five mortality across the considered explanatory variables. The results are presented in Table 2. Occurrence of neonatal mortality was found higher among mothers aged ≥35 years, primary educated mothers, mothers not working, mothers whose husbands were uneducated, mothers who resided in a household whose head was female, mothers who had two or more children at the time of the survey, place of residence was urban, and place of region was Barishal and Rangpur. A similar pattern was observed for infant and under-five mortality, except for the place of region while infant and under-five mortality were found higher in the Sylhet division.

**Table 2:** Distribution of neonatal, infant and under-five mortality across background characteristics, Bangladesh, 2017/18.

| Characteristics | Neonatal mortality<br>per 1000 (95% CI) | Infant mortality<br>per 1000 (95% CI) | Under-five mortality<br>per 1000 (95% CI) |
| --- | --- | --- | --- |
| **Maternal age at birth** |  |  |  |
| ≤19 | 22.8 (16.1-32.1) | 26.0 (18.9-35.5) | 47.4 (37.8-59.2) |
| 20-34 | 21.1 (17.3-25.5) | 27.6 (23.3-32.7) | 38.4 (33.1-44.4) |
| ≥35 | 25.1 (12.6-49.5) | 28.2 (14.7-53.3) | 29.2 (15.6-54.1) |
| **Mother's educational status** |  |  |  |
| No education | 25.6 (15.7-41.6) | 32.1 (20.3-50.4) | 52.6 (35.6-77.1) |
| Primary | 27.2 (20.5-35.9) | 36.0 (28.2-45.7) | 48.1 (39.2-58.9) |
| Secondary | 19.3 (14.9-25.0) | 24.0 (19.1-30.0) | 38.4 (32.3-45.6) |
| Higher | 16.7 (10.0-27.6) | 18.8 (11.7-30.0) | 24.5 (16.3-36.7) |
| **Mother's working status** |  |  |  |
| Yes | 24.1 (19.1-30.3) | 28.9 (23.5-35.5) | 40.1 (35.6-45.2) |
| No | 19.8 (15.87-24.8) | 26.0 (21.4-31.5) | 39.5 (33.9-46.0) |
| **Partner's educational status** |  |  |  |
| No education | 24.8 (16.1-38.0) | 33.8 (23.8-47.7) | 48.9 (36.5-65.3) |
| Primary | 21.2 (16.0-28.0) | 27.2 (21.2-34.9) | 40.1 (32.6-49.1) |
| Secondary | 21.7 (16.1-29.2) | 28.1 (21.6-36.6) | 42.6 (34.5-52.5) |
| Higher | 17.9 (11.6-27.6) | 18.8 (12.4-28.5) | 27.4 (19.7-38.0) |
| **Sex of the household's head** |  |  |  |
| Male | 20.9 (17.6-24.9) | 27.0 (23.2-31.5) | 40.8 (35.9-46.2) |
| Female | 26.2 (17.4-39.3) | 28.7 (19.2-42.5) | 36.1 (25.5-50.9) |
| **Total children ever born** |  |  |  |
| ≤2 | 14.3 (11.2-18.2) | 16.8 (13.6-20.9) | 32.7 (27.8-38.5) |
| >2 | 36.6 (29.2-45.8) | 48.5 (40.0-58.6) | 55.3 (46.0-66.3) |
| **Exposure to mass media** |  |  |  |
| Little exposed | 20.9 (16.3-26.9) | 28.0 (22.4-35.1) | 43.7 (36.0-53.0) |
| Moderate exposed | 14.9 (10.3-29.9) | 20.8 (13.7-31.5) | 39.0 (32.8-46.3) |
| Highly exposed | 24.0 (19.3-29.9) | 28.5 (23.3-34.8) | 35.5 (25.8-48.6) |
| **Wealth quintile** |  |  |  |
| Poorest | 23.2 (17.1-31.3) | 29.5 (22.7-38.3) | 48.4 (38.6-60.5) |
| Poorer | 21.4 (14.9-30.7) | 29.8 (22.2-39.9) | 39.9 (31.0-51.1) |
| Middle | 23.4 (16.7-32.6) | 26.4 (19.3-36.0) | 36.4 (28.1-46.9) |
| Richer | 21.7 (14.5-32.5) | 27.9 (19.4-40.1) | 41.4 (30.7-55.6) |
| Richest | 18.4 (11.9-28.2) | 22.1 (14.8-32.7) | 33.5 (24.4-45.9) |
| **Place of residence** |  |  |  |
| Urban | 25.8 (19.2-34.5) | 30.6 (23.3-40.0) | 43.7 (35.0-54.4) |
| Rural | 20.1 (16.7-24.1) | 26.0 (22.0-30.6) | 38.8 (33.6-44.7) |
| **Place of region** |  |  |  |
| Barishal | 26.1 (16.7-40.6) | 32.0 (22.2-45.9) | 46.1 (34.5-61.2) |
| Chattogram | 23.4 (16.8-32.5) | 25.5 (18.2-35.5) | 36.4 (27.5-48.0) |
| Dhaka | 21.3 (14.7-30.8) | 27.2 (19.5-38.0) | 41.4 (31.3-54.7) |
| Khulna | 19.7 (12.5-30.7) | 25.5 (17.4-37.2) | 34.0 (24.1-47.7) |
| Mymensingh | 12.8 (7.3-22.4) | 17.2 (10.8-27.3) | 34.1 (25.8-44.8) |
| Rajshahi | 17.2 (0.9-30.1) | 24.9 (15.3-40.4) | 42.3 (28.7-62.2) |
| Rangpur | 29.4 (19.5-44.2) | 30.9 (20.8-45.7) | 39.4 (26.7-57.8) |
| Sylhet | 22.7 (14.7-34.8) | 39.2 (29.5-45.7) | 52.3 (40.7-67.1) |

The association of child mortality with health facility level factors were determined using the multilevel mixed-effect logistic regression model. Both unadjusted and adjusted models were run separately for each of the three outcome variables. We found significant effects of healthcare facility level factors on each of the neonatal mortality, infant mortality, and under-five mortality. The associations were mostly persists following the adjustment of the individual-, household-, and community level factors in the adjusted model. The likelihoods of neonatal mortality was found to be declined by 15% (aOR, 0.85; 95% CI, 0.63-0.99) and 16% (aOR, 0.84, 0.62-0.96) for every unit increase of the scores of the basic management and administrative system and the availability of child healthcare services at the nearest healthcare facility, respectively. Similarly, one unit increase in score of the nearest healthcare facility readiness to provide child healthcare services was found to be associated with a 36% (aOR, 0.64; 95% CI, 0.45-0.86) reduction of neonatal mortality. In contrary, for every km increase in distance of the nearest healthcare facility from mothers’ homes where child healthcare services available was found responsible for 1.20 times (95% CI, 1.01-1.45) increase of neonatal mortality.

**Table 3:** Multilevel logistic regression models assessing the relationships between the use of long-acting modern contraception and health facility-, individual-, household-, and community-level factors (N=10,384)

| Characteristics | Neonatal mortality, OR<br>(95% CI) | Infant mortality, OR<br>(95% CI) | Under-five mortality, OR<br>(95% CI) |
| --- | --- | --- | --- |
| **Unadjusted association** |  |  |  |
| Basic management and administrative system | 0.80 (0.60-0.98)** | 0.89 (0.68-1.08) | 0.96 (0.80-1.84) |
| The availability of child healthcare services at the nearest health care facility | 0.63 (0.43-0.85)** | 0.82 (0.68-0.99)** | 0.90 (0.54-1.29) |
| Readiness of the health care facility to provide child healthcare services | 0.64 (0.45-0.86)** | 0.68 (0.38-0.98)** | 0.72 (0.40-0.95)** |
| Average distance on road communication from women's resided cluster to the nearest health facility | 1.28 (1.03-1.55)** | 1.18 (1.01-1.59)** | 1.13 (1.01-1.33)** |
| **Adjusted association<sup>++</sup>** |  |  |  |
| Basic management and administrative system | 0.85 (0.63-0.99)** | 0.89 (0.68-1.08) | 0.95 (0.78-1.86) |
| The availability of child healthcare services at the nearest health care facility | 0.66 (0.45-0.89)** | 0.82 (0.68-0.99)** | 0.93 (0.56-1.30) |
| Readiness of the health care facility to provide child healthcare services | 0.68 (0.47-0.90)** | 0.68 (0.38-0.98)** | 0.75 (0.41-0.96)** |
| Average distance on road communication from women's resided cluster to the nearest health facility | 1.20 (1.01-1.45)** | 1.18 (1.01-1.59)** | 1.15 (1.04-1.36)** |

Notes: \*\* $p < 0.01$ , \* $p < 0.05$ . ++Models are adjusted for maternal age at birth, maternal education, maternal formal employment status, partner's educational status, sex of the household's head, total children ever born, exposure to mass media, wealth quintile, place of residence and region of residence.

Similarly, one unit increase in the scores of the availability of child healthcare services at the nearest health care facility and readiness of the health care facility to provide child healthcare services were found to be associated with a 18% (aOR, 0.82, 95% CI, 0.68-0.99) and 32% (aOR, 0.68, 95% CI, 0.38-0.98) reductions of infant mortality. Moreover, one kilometre increase of the nearest healthcare facility from mothers’ homes was found to be associated with 1.18 times (aOR, 1.18, 95% CI, 1.01-1.59) increase in infant mortality. One unit increase score of the of readiness of the nearest healthcare facility to provide child healthcare services was found associated with a 25% (aOR, 0.75, 95% CI, 0.41-0.96) reduction in under-five mortality. In comparison, under-five mortality was found to be increased by 15% (aOR, 1.15, 95% CI, 1.04-1.36) for every unit increase in the average distance of the nearest healthcare facility from mothers’ homes.

## Discussion

The aim of this study was to explore the associations of several forms of child mortality with healthcare facility level factors. Four healthcare facility level factors were considered which were generated covering management and administrative system of the mothers’ home nearest healthcare facility, availability of the child healthcare services at the mothers’ home nearest healthcare facility, readiness of the mothers’ homes nearest healthcare facility to provide child healthcare services, and average distance of the nearest healthcare facility providing child healthcare services from mothers’ homes. On average, increasing scores of the health facility level factors were found protective to the occurrence of neonatal, infant, and under-five mortality. In addition, around 15-20% increase in the occurrence of neonatal, infant, and child mortality were reported for every kilometre increase in the distance of the nearest healthcare facility providing child healthcare services from mother’s homes. These findings are robust as they were generated by analysing the two nationally representative datasets and adjusted for possible confounders. The findings will help the policymakers to know the area where focus is needed to face ongoing challenges of under-five mortality and in designing policies and programs, accordingly.

There is a common understanding that inadequate healthcare services and poor quality of available services have a negative effect on child health, including child morbidity and mortality [34,35]. However, when this comes to specific dimensions, such as particular area of inadequacy, available research in LMICs are mostly limited and they cannot answer these things. As far as we know, for the first time in LMICs, we considered healthcare facility level factors, including management and administrative system of the mothers’ homes nearest healthcare facility, availability of the child healthcare services at the mothers’ homes nearest healthcare facility and determined their associations with neonatal, infant and under-five mortality. As such we could not validate our findings. However, consideration of such specific wings will help the policymakers to know the areas that need to be prioritized.

In this study, poor administrative and management system, lower availability of the child healthcare services and readiness of the healthcare facility to provide child healthcare services were found to be linked with higher likelihoods of neonatal, infant and under-five mortality. There may be direct and indirect pathways of such associations. Availability of healthcare services and quality of healthcare services in the healthcare centre depend on the proper linkage among several wings of the healthcare facility [36,37]. This includes administrative and management systems, availability of the healthcare personnel and healthcare equipment and their readiness to provide healthcare services [24]. Poor performance in any wing, therefore, affects performance in other wings too, as such indirectly affects the overall performance of the healthcare facility [24]. For instance, the functional availability of the healthcare providing equipment does not always mean its readiness to provide services [38-40]. This may arise because of lack of skilled healthcare personnel to operate equipment and/or lack of proper initiative from the administrative system to ensure use of the equipment and vice versa [40]. This is particularly an issue in Bangladesh as well as other LMICs where healthcare facilities are often run with either inadequate equipment or the healthcare personnel, or both [41,42]. Moreover, development activities in the healthcare facility are often not representative of the community level requirement [43]. This concern is particularly true in the rural area, from where around 75% of the study population were included, where there are no/limited alternative in choosing the healthcare facilities [41].

In Bangladesh, as like other LMICs, governmental healthcare facility plays a driver role in providing child healthcare services, particularly for complicated cases [38,41]. The reasons are: (i) availability of the healthcare facility at lower distance, (ii) free or less expensive healthcare services and (iii) availability of the healthcare personnel and equipment [38,42]. However, healthcare facilities in Bangladesh are not equally equipped and the services available there are not consistent [41]. For instance, healthcare services for complicated cases are only available at the district or divisional level hospitals [11]. Basic services are available at the sub-district, thana and community level hospitals. Consequently, people need to travel to district or divisional level hospitals to access healthcare services in complicated cases for their children. This could increase the occurrence of child mortality as many people do not have the financial capacity to travel to district/divisional level hospitals, buy medicines and stay there for accessing services [44,45]. Even if people have this capacity, transportation is still an important issue considering, (i) transportation to go to the district or divisional level hospitals may not be available particularly for the rural area, and (ii) district or divisional level facilities may stay far and it may take longer hours to go to the healthcare facility [44,45]. This understanding is consistent with this study’s findings of a 15-20% increase in under-five mortality for every kilometre increased healthcare facility nearest to the maternal homes where child healthcare services are available. A similar association has also been reported in other studies in LMICs [44,46,47].

We found effects healthcare facility level factors were strongest for neonatal mortality following the infant and under-five mortality. Moreover, increase scores of the management and administrative system of the mothers’ homes nearest healthcare facilities and services availability of that healthcare facility were found associated with reduced odds of neonatal and infant mortality, but not for the under-five mortality. These associations further support the distance and quality issues of the services are played important roles in the occurrence of under-five mortality. The underlying mechanism could be explained by the children’s immune power which is increased with the increased age of children [48]. As such, children who are comparatively high at age have usually better immune power. Consequently, hospital admission is comparatively lower among higher-aged children as compared to the newborn because they mostly get recovered from diseases with minimal treatments available at the community level [49]. Even for complicated cases, such better immune power of the comparatively higher aged children keep them survive for a moderately higher time, as such their parents geta chance to bring them to the healthcare facilities, if even it is located in far [45]. An opposite direction is reported for neonatal and infant mortality as they have comparatively poor immune power, therefore parents may not get enough time to bring them to the healthcare facility.

The implications of our findings are that there needs to be substantial investment in the healthcare sector to ensure availability of the child healthcare services at the community level healthcare facilities, such as community clinic. However, healthcare facilities where child healthcare services are available are usually located in urban areas and rural healthcare facilities are often under-resourced and are not capable of providing the child healthcare services. The reasons are lack of skilled healthcare personnel at the community level healthcare facilities, poor or under-developed infrastructure and lack of medical equipment. Unchanged government budget allocations in healthcare sectors over the years (2.02% [2000] and 2.37% [2016] of gross domestic product) in the face of rising healthcare demands are often causes of such crisis [50]. Transportation services in the urban healthcare facilities to bring children who need critical care to the urban healthcare facilities are also lacking. Therefore, priority also needs to be given to improving the transportation facilities of the healthcare facilities. This needs universal healthcare coverage-a target that is also present in the SDGs to be achieved by 2030. Consequently, the focus is needed on the governmental level policies and programs.

This study has several strengths and some limitations. The most important strength of this study is the analysis of two nationally representative household and healthcare facility surveys data. By this, the associations of healthcare facility level factors with several form of child mortality were determined. As far as we know this sort of understanding is first in Bangladesh and LMICs. Moreover, findings were adjusted with the range of adjustment factors at the individual-, households-, and community-level factors. Advanced statistical modelling was used to determine the associations. Therefore, the findings of this study are robust and could be applicable for the national level policy and program making. However, since we analysed a cross-sectional household survey data, the findings is correlational only, not casual. To secure the privacy of the respondents, BDHS displaced clusters’ locations up 0-5 km for rural areas and 0-2 km for urban areas. Therefore, the calculated average distance of the nearest health facility could be slightly different from the actual distance. However, the BDHS ensured that the new perturbed locations fell within the designated administrative boundaries. Therefore, errors from displacement are likely to be random and minimum. A previous study found that the effect of this variation is insignificant [51]. Regardless of these limitations, to our knowledge, this is the first study in the context of LMICs and Bangladesh that examined the influence of health facility-level factors on under-five mortality adjusted for individual-, household-, and community-level factors. Therefore, the finding of this study is likely to be precise and can be generalisable in countries with similar features to Bangladesh and may help in making evidence-based policies.

## Conclusion

We reported prevalence of under-five, infant and neonatal mortality rates in Bangladesh were 40, 27 and 21 per 1000 live births. Increase scores on the basic management and administrative system were found negatively associated with the occurrence of neonatal mortality. In addition, availability of child healthcare services at the nearest healthcare facility and readiness of the mothers’ homes nearest healthcare facility to provide child healthcare services were found negatively associated with neonatal and infant mortality. Average distance on road communication from mothers’ resided clusters to the nearest health facility was found to be linked with the 15-20% increase in the likelihoods of neonatal, infant and under-five mortality. Healthcare facilities should be strengthened to provide child healthcare services, particularly in the rural area where transportation is tan ongoing issue. Healthcare facilities’ transportation facilities, including ambulance services, should also be strengthened.

## Data Availability

The data that support the findings of this study are available from The DHS Program, but restrictions apply to the availability of these data, which were used under license for the current study, and so are not publicly available. Data are, however, available from the authors upon reasonable request and with permission of The DHS Program. Data are available here: https://dhsprogram.com/data/available-datasets.cfm

## Declarations

## Acknowledgements

We acknowledge the support of Research and Extension Centre, Jatiya Kabi Kazi Nazrul Islam University.

## Ethics approval and consent to participate

The survey analysed was approved by the institutional review board of ICF and the National Research Ethics Committee of the Bangladesh Medical Research Council. Informed consent was obtained from all participants. All necessary patient/participant consent has been obtained and the appropriate institutional forms have been archived. No separate ethical approval was required to conduct this study. We obtained permission to access this survey and conduct this research. All methods were performed in accordance with the relevant guidelines and regulations.

## Authors’ contributions

MNK designed the study, performed the data analysis, and wrote the first draft of this manuscript. NIT and MMR critically reviewed and edited all versions of this manuscript. All authors approved this final version of the manuscript.

## Consent for publication

Informed consent was obtained from all participants during the survey using standard DHS consent form.

## Availability of data and materials

The data that support the findings of this study are available from The DHS Program, but restrictions apply to the availability of these data, which were used under license for the current study, and so are not publicly available. Data are, however, available from the authors upon reasonable request and with permission of The DHS Program.

